# Differential microRNA expression profiling of peripheral blood L1CAM neural-enriched and bulk extracellular vesicles in individuals with bipolar disorder

**DOI:** 10.1101/2025.06.06.25329080

**Authors:** Gabriel R. Fries, Salahudeen Mirza, Jun Wang, Camila N. C. Lima, Wei Zhang, Marcela Carbajal Tamez, Giselli Scaini, Jair C. Soares, Joao Quevedo

## Abstract

Bipolar disorder (BD) is a common and yet poorly elucidated psychiatric disorder, with emerging evidence implicating a role for epigenetic mechanisms, including microRNAs (miRNAs), in its pathophysiology. These molecules are secreted from cells in extracellular vesicles (EVs), which can be isolated from bodily fluids and tested as potential biomarkers. In individuals with BD and control participants (CON), we characterized the miRNA expression profiles of peripheral blood EVs selected for L1CAM, a putative neuronal marker, as well as bulk peripheral blood EVs. Peripheral blood EVs were isolated from n=20 BD and 20 CON (L1CAM) and n=21 BD and 20 CON (bulk). Within each study, analyses identified miRNAs that were differentially expressed between BD and CON, followed by functional interrogation and testing for associations with clinical features. Results were then compared to better understand the relative specificity of bulk and L1CAM EV analyses. Thirty-four miRNAs were differentially expressed between groups in L1CAM EVs, whereas 10 differentially expressed miRNAs were identified in bulk EVs. Across both analyses, biological pathways attributed to the differentially expressed miRNAs included insulin receptor pathway and type II diabetes mellitus. Importantly, associations of differentially expressed miRNAs with clinical features were only significant in L1CAM EVs. Our results reiterate a crucial role for miRNAs in the pathophysiology of BD and suggest that miRNA signatures of putative neuronal origin more closely correspond to clinical features.

## 1. Introduction

Bipolar disorder (BD) is a potentially severe and highly prevalent psychiatric disorder characterized by pronounced mood and energy disturbances of (hypo)manic and depressive episodes (Carvalho et al. 2020). Beyond the distress associated with its symptoms, BD is also associated with other significant risks, such as high rates of psychiatric comorbidity, increased likelihood of other medical conditions and premature death, cognitive and functional impairments, and an elevated risk for suicide compared to the general population (Carvalho et al. 2020).

BD is presumed to have a strong biological basis (Legrand et al., 2021; Scaini et al., 2020). Early genetic studies demonstrated a strong heritability for BD when considering familial aggregations and patterns in mono- and dizygotic twins. More recently, several gene expression changes in the brain have been documented, providing a possible molecular basis for the pathology of BD (Seiffudin et al., 2013). At the same time, genome-wide association studies of common variants have only partially been able to account for the original heritability from genetic epidemiology studies (O’Connell et al. 2025), possibly suggesting the contribution of other factors.

Epigenetic modulation of gene expression involves both genetic and experiential influences. Epigenetic changes, which include those that operate at the transcriptional, post-transcriptional, translational, and post-translational levels of gene and protein expression, regulate gene expression and can persist across cell division cycles without changing the DNA sequence. There is a growing interest in the possibility that epigenetic alterations play a key role in the pathophysiology of BD (Fries et al. 2016; Legrand et al., 2021; Lima et al., 2021), considering that they may mediate both the effects of genetic influences and environmental exposures, such as early life stress.

MicroRNAs (miRNAs) are short (∼20 nucleotide) non-coding RNAs which bind the miRNA seed - typically the 3’-untranslated segment of messenger RNAs (mRNAs) - to direct downregulation of gene expression by RNA-induced silencing complex (RISC) (Cherone et al. 2019). There are hundreds of distinct miRNAs (Friedman et al. 2009), and miRNAs may be involved in the regulation of expression of 30% of all genes (Kapplingattu et al. 2025). One miRNA may regulate the activity of hundreds of target mRNAs, suggesting a critical master regulatory role. Several miRNAs have been shown to be differentially expressed in BD (Fries et al. 2018), though convergence across studies – even within tissue type - is limited (Martinez & Peplow, 2024). Though miRNA expression can be and has been tested in peripheral blood, the correlation of these findings with miRNA expression changes in the brain is unclear. The present inability to directly assess brain miRNA expression *in vivo* restricts analyses of brain tissue to the post-mortem context, which introduces new sources of ambiguity and confounding.

Extracellular miRNAs have been detected in various biological fluids, including blood (O’Brien et al., 2018). A subset of miRNAs is present in extracellular vesicles (EVs), such as exosomes, micro-vesicles, and apoptotic bodies (Fries & Quevedo, 2018). These miRNAs are important mediators of intercellular communication, but they also can serve as biomarkers of disease. Given that EVs carry molecular markers that identify their cellular origin, they can be leveraged to better understand miRNA expression patterns in otherwise inaccessible cell populations, such as neurons. Selecting EVs for neuronal origin may provide insights to brain miRNA expression profiles in living patients, circumventing challenges associated with interpreting miRNA expression patterns in post-mortem human brain tissues.

EVs selected in blood for L1 cell adhesion molecule (L1CAM) show a strong signal suggestive of neuronal origin, despite some controversy (Nogueras-Ortiz et al., 2024). Supporting evidence demonstrates the correlation of L1CAM EV contents with brain pathology (Delgado-Peraza et al., 2021; Mullins et al., 2017; Walker et al., 2021), with studies showing that L1CAM EVs also show brain-specific neuronal markers and contain miRNAs enriched for brain functions (Saeedi et al., 2021). To our knowledge, miRNA expression profiles associated with BD have not been compared between L1CAM EVs and bulk EVs, limiting our understanding of the brain-specific miRNA expression profiles of living individuals with BD.

We carried out two analyses to better understand miRNA expression patterns in BD. In the first one, we assessed miRNA expression differences between individuals with BD and non-psychiatric controls in L1CAM EVs from peripheral blood. We then used an independent cohort to assess miRNA expression differences between individuals with BD and non-psychiatric controls in bulk EVs from peripheral blood. We hypothesized that miRNA expression differences between BD and non-psychiatric controls may be more pronounced in the L1CAM EVs, and furthermore that the implicated biological pathways would be more specific to brain development and function. We also hypothesized that miRNAs implicated in the L1CAM EVs may be more specifically related to clinical features of BD, again suggesting a greater brain specificity and potential future clinical applicability.

## 2. Methods

### 2.1. Subjects

Participants were recruited at the Center of Excellence in Mood Disorders, McGovern Medical School, University of Texas Health Science Center at Houston, Houston, TX. The Structured Clinical Interview for DSM-IV (SCID-IV) was used for the diagnosis of BD, and individuals with BD were not required to be euthymic at recruitment. Also according to the SCID-IV, participants were admitted to the non-psychiatric control group if they had no lifetime or present history of any major psychiatric disorder or such a history in any first-degree relatives. For the analysis of L1CAM peripheral blood EVs, n = 20 individuals with BD and n = 20 non-psychiatric controls were recruited. For the analysis of bulk peripheral blood EVs, n = 21 individuals with BD and n = 20 non-psychiatric controls were recruited.

In both cohorts, the severity of current depressive and manic symptoms in individuals with BD was assessed using the Montgomery-Åsberg Depression Rating Scale (MADRS) (Williams and Kobak 2008) and the Young Mania Rating Scale (YMRS) (Young et al. 1978), respectively. Functional status was assessed in individuals with BD and non-psychiatric controls with the Global Assessment of Functioning (GAF) (Aas 2011) and the Functioning Assessment Short Test (FAST) (Rosa et al. 2007). Impulsivity was assessed with the Barratt Impulsivity Scale (BIS) (Rosa et al. 2007; Barratt 1965), and the Hollingshead Four-Factor Index of Socioeconomic Status (SES-Adult) (Cirino et al. 2002) was used to assess the socioeconomic status of all participants.

All procedures were performed in compliance with relevant laws, and the Institutional Review Board (IRB) approved the overall study at the University of Texas Health Science Center at Houston (IRB number HSC-MS-19-0157, 3/27/2019). All participants provided a signed informed consent after receiving a full explanation of the study procedures, and their privacy rights have been observed.

### 2.2. EV isolation, immunoprecipitation, and characterization

Fasting peripheral blood was collected from all subjects into heparin-containing vacutainers, followed by separation and storage of plasma in -80°C freezers. EVs were then isolated from all plasma samples with the ExoQuick® ULTRA EV Isolation Kit (Systems Biosciences). In brief, 1mL plasma samples from all subjects were initially centrifuged at 3,000 × g for 15 minutes at 4°C to remove cellular debris, followed by treatment of the supernatant with 5U thrombin. After a 5-minute incubation at room temperature, the fibrin was cleared with a second spin at 10,000 x g for 5 minutes and the resulting supernatant was transferred to a fresh tube for EV precipitation by the ExoQuick® ULTRA EV Isolation Kit, according to the manufacturer’s instructions.

For L1CAM-specific analysis, the samples were further subjected to immunoprecipitation for the L1CAM neural adhesion protein, a putative marker for neural-enriched EVs (Dunlop, Banack, and Cox 2023; Gomes and Witwer 2022; Serpente et al. 2020), using the mouse anti-human CD171 (L1 cell adhesion molecule [L1CAM]) biotinylated antibody (CD171, clone 5G3, eBioscience). Specifically, the EV samples were incubated with this antibody at 4°C for 2 hours on a rotating mixer and then precipitated using the Pierce streptavidin-agarose Ultralink resin (Thermo Scientific, Rockford, IL, USA) for 4 hours at 4°C. After centrifugation at 200 x g at 4°C for 10 minutes, the pellet containing neural-enriched EVs was washed twice with PBS and stored at -80°C. Finally, the isolated L1CAM EVs were characterized by Western blot for the presence of common EVs and neural-specific markers, specifically: NCAM-L1 (C-2, sc-514360, Santa Cruz Biotechnology), microtubule-associated protein 2 (MAP2 (C-2), sc-390543, Santa Cruz Biotechnology), N-enolase (clone 5E2, MAB324, EMD Millipore), neuron-specific class III beta-tubulin (TUJ1, ab18207, Abcam), CD9 (H-110, sc-9148, Santa Cruz Biotechnology), and tumor suppressor TSG1 (TSG1, ab133586, Abcam).

### 2.3. RNA-sequencing

In both cohorts, total RNA was isolated from the bulk EVs and L1CAM-enriched EVs with the exoRNeasy Midi Kit (Qiagen) and Seramir RNA Isolation Kit (Systems Biosciences), respectively, according to the manufacturer’s instructions. RNA quality was checked on Bioanalyzer (Agilent) using the Agilent RNA 6000 Pico Kit and assessed for the presence of miRNAs. For the study of L1CAM EVs, total isolated RNA was entered into a small RNA library preparation using Seqmatic’s TailorMix for paired-end sequencing (SeqMatic). For the analysis of bulk EVs, the total RNA population, starting with as little as 1 ng of total RNA, was entered into small RNA library prep using Qiagen’s miRNA library kit for single-end sequencing (Qiagen). In both analyses, libraries were amplified by qPCR and assessed for viability by the concentration (> 1nM) to be moved on to sequencing. Successful libraries were purified by gel extraction to ensure the correct size was isolated, and libraries were sequenced on the Illumina NextSeq using a small RNA-Seq (Illumina Platform), 1x51bp, 10M (avg) raw reads per sample (single-end). To obtain read counts per miRNA per sample, sequencing reads were submitted to the ERCC exceRpt small RNA-seq pipeline (v.5.0.0) with default settings (adapter trimming, reads filtering, alignment to the human genome GRCh38 according to miRNA annotation in the miRBase v22). After filtering steps, 165 miRNAs were detected in L1CAM EVs and 188 miRNAs were detected in bulk EVs for differential expression (DE) analysis with DESeq2 (v.1.30.1). Raw RNA-sequencing data and accompanying phenotype information are deposited at the NIMH Data Archive (NDA, collection ID #3300).

### 2.4. Differential expression analyses

First, for each cohort, demographic characteristics of the BD and non-psychiatric control groups were compared using chi-square for categorical variables and Mann-Whitney U tests for continuous variables. In each analysis, models comparing miRNA expression between individuals with BD and non-psychiatric controls were adjusted for age (median split) and sex. Benjamini-Hochberg false discovery rate (FDR) correction was applied with significance threshold of Q < .10.

### 2.5. Pathway over-representation analyses

FDR-significant differentially expressed miRNAs were input to the miEAA 2.1 web tool (Aparicio-Puerta et al. 2023) to identify enrichment for gene ontology (GO) processes (miRTarBase, miRWalk), diseases (MNDR), pathways (KEGG, miRWalk), and target genes (miRTarBase).

### 2.6. Classification analyses

For the top-ranked differentially expressed miRNAs in each analysis, the association between the normalized expression (by variance stabilizing transformation from DESeq2) of miRNAs and the classification of BD *vs.* controls was further evaluated with logistic regressions. miRNAs were tested independently, and the area under the ROC curve (AUC) values were calculated with the *caret* (v6.0.93) (Kuhn 2008), *AUC* (v0.3.2), and *cvAUC* (v1.1.4) R packages.

### 2.7. Association of miRNA expression with clinical features

Among individuals with BD, the association of expression level of each of the significant (Q < .10) miRNAs from the main analysis was tested using Spearman or Pearson correlations for the following clinical variable: age at diagnosis of any mood disorder; age at diagnosis of BD; age at first mood stabilizer treatment; number of manic, hypomanic, mixed, depressive, and total episodes; number of prior hospitalizations; MADRS scores; YMRS scores; and FAST scores. Significant correlations were identified if Q < .05.

## 3. Results

### 3.1 Samples

For both cohorts, there were no significant differences between individuals with BD and controls on sex, age, race/ethnicity, or body mass index (**Table 1**). However, individuals with BD had lower socio-economic status, lower functioning, and higher impulsivity.

**Table 1.**
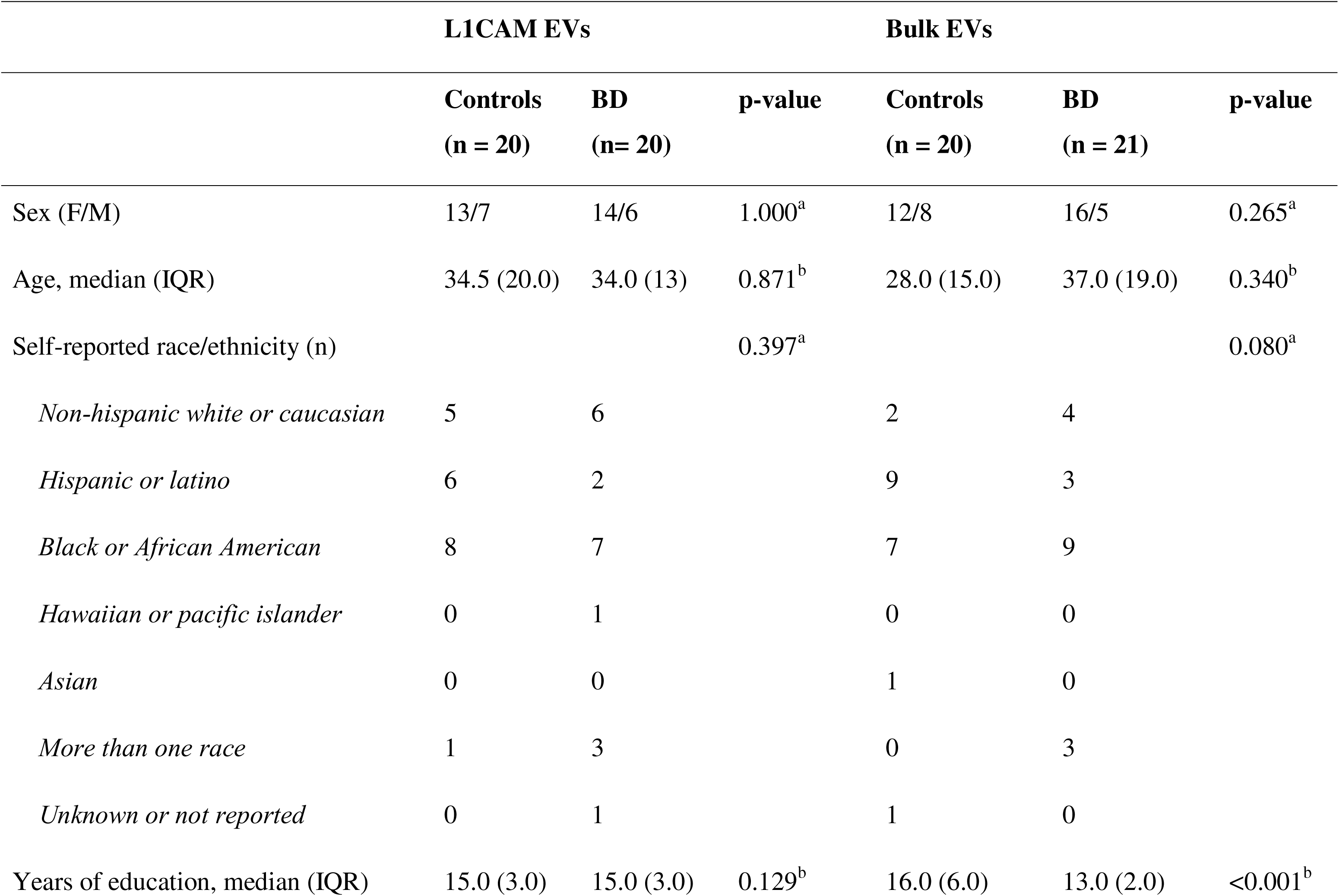
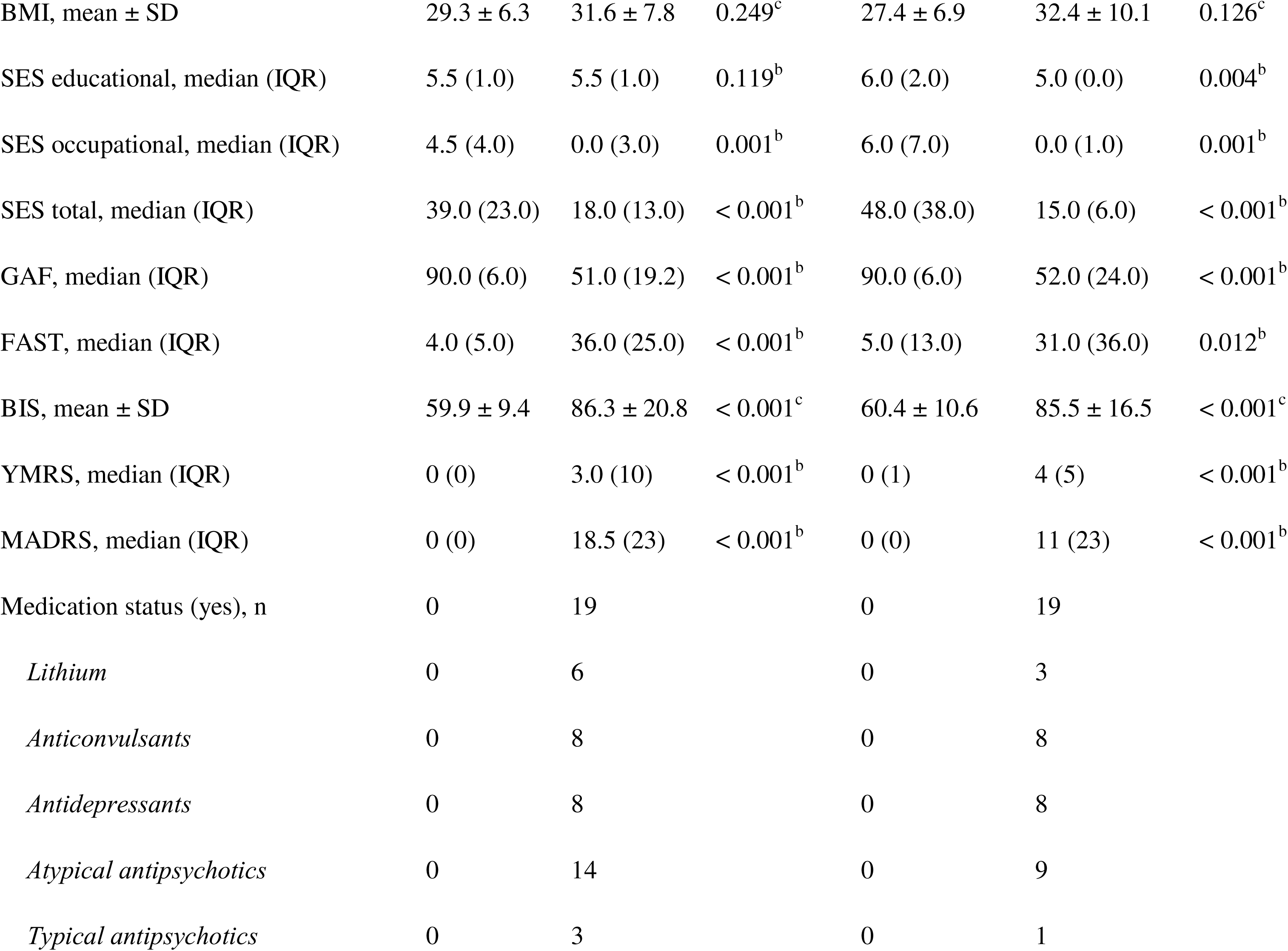
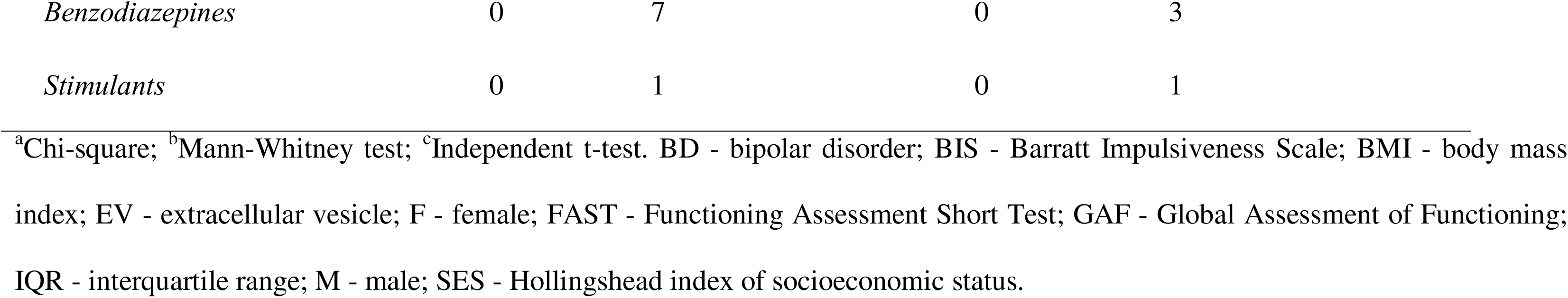
Sample demographics.

### 3.2 L1CAM EVs

#### 3.2.1 Immunoprecipitation for L1CAM

Western blot suggested the successful enrichment of EVs for neural origin using the L1CAM marker (**Supplementary Figure 1**), as suggested by an increase in the levels of the neural-specific markers MAP2, N-enolase, and TUJ1 after immunoprecipitation compared to total EVs.

#### 3.2.2 Differential miRNA expression and pathway overrepresentation analyses

Thirty-four miRNAs were significantly differentially expressed between individuals with BD and non-psychiatric controls at the significance threshold of Q <.10 in the L1CAM EVs (**Figure 1A and Table 2**). The top-ranked differentially expressed miRNAs include hsa-miR-486-5p, hsa-miR-486-3p, hsa-miR-128-3p, and hsa-miR-301a-3p. Follow-up analyses implicated *PTEN*, *TMED5*, and *EZH2* as the top-ranked target genes (**Supplementary Figure 2A**), with significant enrichment for multiple signaling pathways (**Supplementary Figure 3**). Finally, as exploratory analyses, we also found the ten top-ranked miRNAs to have high discriminatory accuracy, with classification of BD *vs.* non-psychiatric control ranging from 91.2% for hsa-miR-486-5p to 55% for hsa-miR-150-5p (**Supplementary Figure 4**).

**Figure 1.**
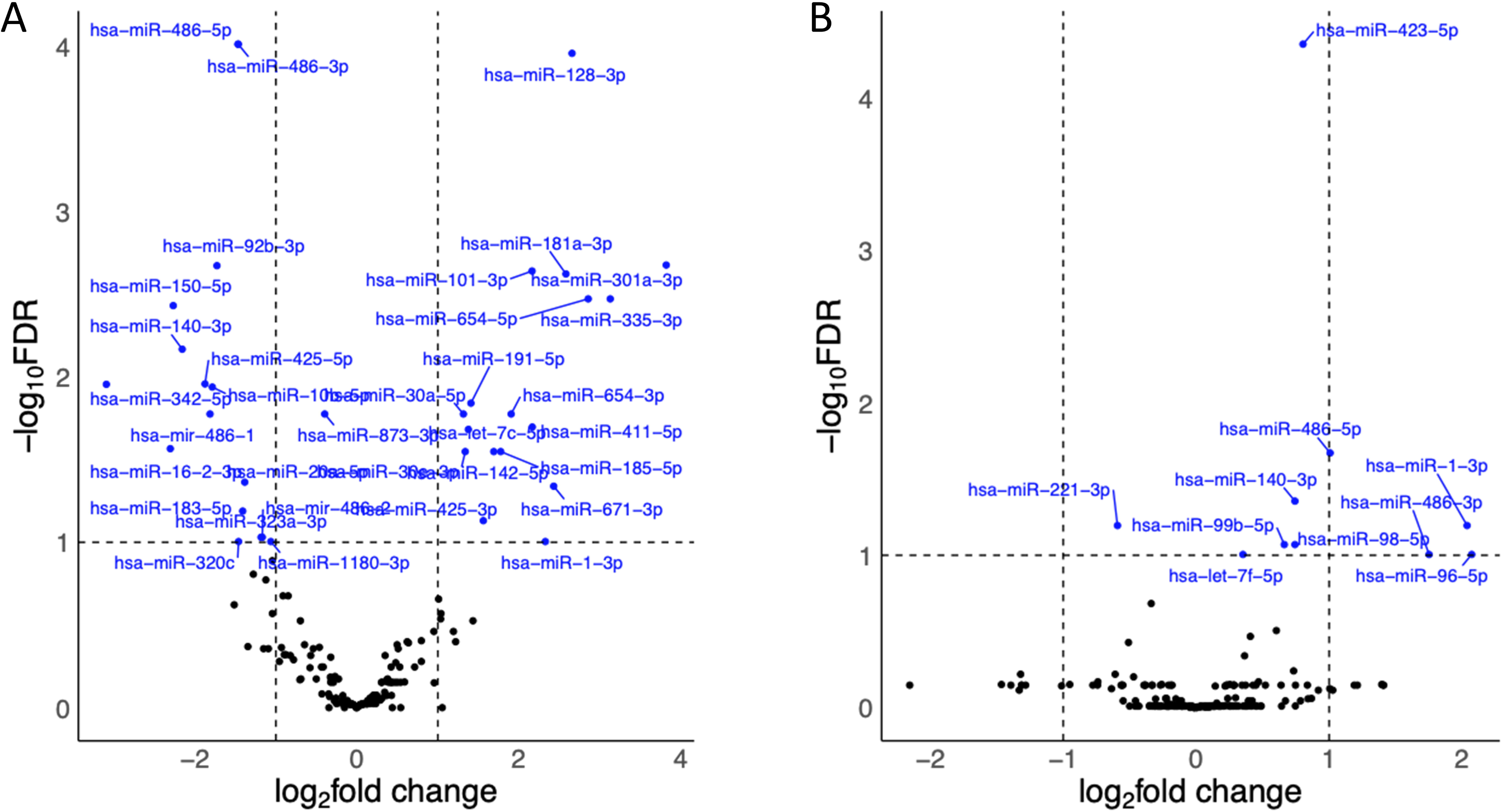
Volcano plots showing differentially expressed microRNAs in individuals with bipolar disorder compared to controls. Blue dots indicate those with a false discovery rate-corrected p < 0.1. A) L1CAM extracellular vesicles; B) Bulk extracellular vesicles.

**Table 2.** Differentially expressed microRNAs in L1CAM EVs (FDR < 0.1).

| miRNA | log2FoldChange | lfcSE | stat | P-value | Adjusted P-value |
| --- | --- | --- | --- | --- | --- |
| hsa-miR-486-5p | -1.4669665 | 0.29570203 | 23.6881651 | 1.13E-06 | 9.73E-05 |
| hsa-miR-486-3p | -1.4651403 | 0.29614689 | 23.5636594 | 1.21E-06 | 9.73E-05 |
| hsa-miR-128-3p | 2.65680457 | 0.52314647 | 22.5392913 | 2.06E-06 | 0.00011049 |
| hsa-miR-301a-3p | 3.819899 | 0.67559144 | 16.3617044 | 5.23E-05 | 0.00210636 |
| hsa-miR-92b-3p | -1.7274803 | 0.4164706 | 15.9222826 | 6.60E-05 | 0.00212511 |
| hsa-miR-101-3p | 2.16332003 | 0.51207485 | 15.4368256 | 8.53E-05 | 0.00228914 |
| hsa-miR-181a-3p | 2.58137408 | 0.61765982 | 15.0690982 | 0.00010365 | 0.00238386 |
| hsa-miR-654-5p | 2.85613228 | 0.5932544 | 13.9416686 | 0.00018857 | 0.00337334 |
| hsa-miR-335-3p | 3.12853461 | 0.61043971 | 14.0898111 | 0.00017429 | 0.00337334 |
| hsa-miR-150-5p | -2.2672009 | 0.56028808 | 13.5675432 | 0.00023013 | 0.0037051 |
| hsa-miR-140-3p | -2.1562699 | 0.56086746 | 12.2527995 | 0.00046456 | 0.00679948 |
| hsa-miR-425-5p | -1.874957 | 0.52805481 | 11.1912783 | 0.00082183 | 0.01102618 |
| hsa-miR-342-5p | -3.0930051 | 0.76830545 | 11.0325751 | 0.00089525 | 0.01108729 |
| hsa-miR-10b-5p | -1.7847615 | 0.50877097 | 10.8240953 | 0.00100188 | 0.01152158 |
| hsa-miR-191-5p | 1.40817338 | 0.42232255 | 10.2827236 | 0.00134282 | 0.01441289 |
| hsa-miR-30a-5p | 1.31521626 | 0.41278204 | 9.75031122 | 0.00179292 | 0.0167611 |
| hsa-mir-486-1 | -1.8130077 | 0.54676675 | 9.57487965 | 0.00197258 | 0.0167611 |
| hsa-miR-654-3p | 1.90531444 | 0.58282119 | 9.56982373 | 0.00197802 | 0.0167611 |
| hsa-miR-873-3p | -0.3966612 | 2.38227865 | 9.70499728 | 0.00183768 | 0.0167611 |
| hsa-miR-411-5p | 2.16616972 | 0.58611893 | 9.14337755 | 0.0024962 | 0.0200944 |
| hsa-let-7c-5p | 1.37637636 | 0.44650939 | 8.99167374 | 0.00271212 | 0.02079296 |
| hsa-miR-16-2-3p | -2.3034697 | 0.6497919 | 8.42017167 | 0.00371081 | 0.02715636 |
| hsa-miR-142-5p | 1.6905999 | 0.55103145 | 8.24177495 | 0.00409371 | 0.02828299 |
| hsa-miR-30e-3p | 1.33823837 | 0.44574262 | 8.14178118 | 0.00432568 | 0.02828299 |
| hsa-miR-185-5p | 1.77606561 | 0.55822698 | 8.11428963 | 0.00439177 | 0.02828299 |
| hsa-miR-20a-5p | -1.3842146 | 0.48272539 | 7.27507358 | 0.0069918 | 0.04329539 |
| hsa-miR-671-3p | 2.43044774 | 0.69851033 | 7.10620185 | 0.00768177 | 0.04580611 |
| hsa-miR-183-5p | -1.4094638 | 0.50094609 | 6.42295147 | 0.01126548 | 0.0647765 |
| hsa-miR-425-3p | 1.56170518 | 0.60595642 | 6.12487984 | 0.01332923 | 0.07400023 |
| hsa-mir-486-2 | -1.1677503 | 0.47265926 | 5.60128815 | 0.01794728 | 0.09321005 |
| hsa-miR-323a-3p | -1.1827989 | 0.47100062 | 5.62386967 | 0.01771749 | 0.09321005 |
| hsa-miR-1-3p | 2.32711194 | 0.6865979 | 5.33504407 | 0.02090081 | 0.09897149 |
| hsa-miR-320c | -1.4606309 | 0.60332152 | 5.42303744 | 0.01987276 | 0.09897149 |
| hsa-miR-1180-3p | -1.0621835 | 0.4563638 | 5.3539599 | 0.02067526 | 0.09897149 |

FDR - false discovery rate; SE – standard error.

#### 3.2.3 Association of significant miRNAs with clinical features in individuals with BD

Two of the significantly differentially expressed miRNAs in L1CAM EVs were significantly associated with clinical features among the individuals with BD only (FDR q < 0.1). Expression level of hsa_miR_92b_3p (downregulated in BD) was negatively associated with the number of previous manic episodes (Spearman’s rho = -0.744, q = 0.007). Expression level of hsa_miR_101_3p (upregulated in BD) was negatively associated with age at first mood stabilizer treatment (rho = -0.585, q = 0.098) and positively associated with total MADRS score (rho = 0.688, q = 0.035).

### 3.3 Bulk EVs

#### 3.3.1 Differential miRNA expression and pathway overrepresentation analyses

Ten miRNAs were significantly differentially expressed between individuals with BD and non-psychiatric controls at the significance threshold of Q < .10 in the bulk EVs (**Figure 1B and Table 3**). The top-ranked differentially expressed miRNAs include the hsa-miR-423-5p, hsa-miR-486-5p, and the hsa-miR-140-3p. Follow-up analyses implicated *ACER2*, *UST*, and *ATP6V1E1* as the top-ranked target genes (**Supplementary Figure 2B**), with significant enrichment for multiple signaling pathways (**Supplementary Figure 5**). As exploratory analyses, we also found all identified bulk EV miRNAs to have high discriminatory accuracy, with classification of BD *vs.* non-psychiatric control ranging from 88.7% for hsa-miR-423-5p to 63% for hsa-miR-1-3p (**Supplementary Figure 6**).

**Table 3.** Differentially expressed microRNAs in bulk EVs (FDR < 0.1).

| **miRNA** | **log2FoldChange** | **lfcSE** | **stat** | **P-value** | **Adjusted P-value** |
| --- | --- | --- | --- | --- | --- |
| hsa-miR-423-5p | 0.80352464 | 0.15439345 | 26.705456 | 2.37E-07 | 4.43E-05 |
| hsa-miR-486-5p | 1.00710602 | 0.27418755 | 13.5853845 | 0.00022795 | 0.02131364 |
| hsa-miR-140-3p | 0.74282238 | 0.21685028 | 11.4685759 | 0.00070783 | 0.04412134 |
| hsa-miR-221-3p | -0.5920304 | 0.18812329 | 9.84134791 | 0.00170632 | 0.06381652 |
| hsa-miR-1-3p | 2.03756803 | 0.50103404 | 10.0028933 | 0.00156294 | 0.06381652 |
| hsa-miR-99b-5p | 0.66248958 | 0.21805435 | 8.96678646 | 0.00274932 | 0.08507664 |
| hsa-miR-98-5p | 0.74285555 | 0.25125364 | 8.69851886 | 0.00318469 | 0.08507664 |
| hsa-let-7f-5p | 0.35174182 | 0.12596727 | 7.77959259 | 0.00528397 | 0.09881026 |
| hsa-miR-96-5p | 2.0721563 | 0.69002176 | 7.83458483 | 0.00512559 | 0.09881026 |
| hsa-miR-486-3p | 1.75176683 | 0.60948852 | 7.8437722 | 0.00509961 | 0.09881026 |

FDR - false discovery rate; SE – standard error.

#### 3.3.2 Association of significant miRNAs with clinical features in individuals with BD

None of the significantly differentially expressed miRNAs in bulk EVs were significantly associated with clinical features among the individuals with BD only (FDR q < 0.1).

### 3.4 Comparison of L1CAM and bulk EV results

Four miRNAs were significantly (Q < .10) differentially expressed in both L1CAM and bulk EVs: hsa-miR-486-5p, hsa-miR-140-3p, hsa-miR-1-3p, and hsa-miR-486-3p. Only hsa-miR-1-3p had the same direction of change across both types of EVs (**Supplementary Table 1**). Also, *ITGA6* emerged as a common gene target in both sub-studies.

## 4. Discussion

In this set of two parallel analyses, we characterized peripheral blood for miRNA expression in EVs of putative neuronal origin and all sources in individuals with BD and control participants. We validated that our L1CAM EVs showed markers consistent with neuronal origin, providing a possible ‘liquid biopsy’ of brain miRNA expression. Our study allowed us to compare the results of this targeted approach to a less restrictive, bulk approach and the ensuing impact on the identified expression differences. In both analyses, several miRNAs were significantly differentially expressed between BD and non-psychiatric controls, with preliminary evidence suggesting that focusing on EVs of presumed neural origin may lead to more clinically significant findings.

Interestingly, L1CAM EVs returned a larger number of differentially expressed miRNAs as compared to bulk EVs, though whether this reflects a stronger signal as opposed to an effect of a lower burden of multiple comparisons correction due to a smaller number of miRNAs considered is unclear. Nevertheless, in line with the idea that the L1CAM EVs may more closely capture brain pathology associated with BD was the finding that expression levels of the significantly differentially expressed miRNAs in L1CAM, but not bulk EVs, were associated with clinical features of BD. We also found that the analysis of L1CAM EVs identified differentially expressed miRNAs with overall higher fold changes between groups compared to bulk EVs (mean absolute log fold change of 1.89 and 1.07 for L1CAM and bulk EVs, respectively), as well as higher discriminatory abilities.

All but six of the 35 differentially expressed miRNAs identified in the L1CAM EVs were part of the bulk EV dataset used as input for the differential expression analysis. However, the only miRNA that was significantly differentially expressed in both sub-studies with the same direction of change was miR-1-3p, which was increased in individuals with BD. Interestingly, miR-1-3p has been shown to target the brain-derived neurotrophic factor (*BDNF*) gene and suppress its expression (Gao et al. 2018), with evidence of a negative correlation between miR-1-3p expression and serum BDNF levels in clinical samples (Ding et al. 2022). BDNF has been repeatedly found to be downregulated in individuals with BD, particularly during acute mood episodes (Fernandes et al. 2015; Munkholm, Vinberg, and Kessing 2016), and our results point to a possible miR-1-3p-mediated mechanism driving this downregulation. Of note, miR-30a-5p, which was increased in BD in L1CAM EVs, can also target and reduce the expression of BDNF (Darcq et al. 2015; Mellios et al. 2008), suggesting that many miRNAs could be acting in synergy to reduce BDNF levels in BD.

We also explored pathways that were significantly overrepresented when considering the significantly differentially expressed miRNAs’ gene targets. Overall, seventeen pathways (KEGG) were shared between miRNAs identified in L1CAM EVs and bulk EVs, including many pathways known to be associated with BD, such as type II diabetes mellitus (Calkin et al. 2013), folate biosynthesis (Hsieh et al. 2019), Th17 cells (Poletti, Mazza, and Benedetti 2024; Poletti et al. 2017), and Alzheimer’s disease (Y. Liu et al. 2024; Liou et al. 2023; Drange et al. 2019). Interestingly, the top-ranked KEGG pathway enriched with the L1CAM EVs miRNAs was insulin signaling pathway, which has been previously linked to BD’s pathology (Calkin et al. 2021) and further suggested as a therapeutic target of the mood stabilizer lithium (Campbell et al. 2022). This finding is also supported by a previous report showing an association between insulin receptor substrate 1 (IRS-1) phosphorylation at serine site 312 (an indicator of insulin resistance) measured in neural-derived EVs and cognitive dysfunction in individuals with BD (Mansur et al. 2021). Overall, our study suggests that miRNAs in the brain may contribute to insulin resistance in BD, proposing them as potential therapeutic targets for future studies.

To our knowledge, this is the first study of L1CAM EV miRNA expression in BD to date. Our group previously found no significantly differentially expressed bulk EV miRNAs between individuals with BD and controls (n=41) after adjustment for multiple comparisons, and we proceeded with exploring the 33 nominally-significant miRNAs instead (Fries et al. 2019). Using a larger sample size (n=110), Ceylan et al. (2020) identified four differentially expressed miRNAs in bulk EVs between individuals with BD and controls after controlling for multiple comparisons (Ceylan et al. 2020). Among the differentially expressed miRNAs identified in this study, only the miR-185-5p (L1CAM EVs) has been previously found in bulk EVs from individuals with BD (Ceylan et al. 2020). Encouragingly, our study also replicated the previously identified increase in miR-185-5p levels in BD compared to controls. This miRNA, which has also been found to be downregulated in peripheral blood of individuals with major depressive disorder (Żurawek and Turecki 2021), has been linked to multiple pathways related to cholesterol homeostasis (Żurawek and Turecki 2021; Tan et al. 2022), mammalian target of rapamycin complex (mTORC) regulation (Chen et al. 2017), and beta-cell proliferation (Bao et al. 2015), all of which are mechanisms previously linked to BD (Guidara et al. 2021; Vanderplow et al. 2021; Charles, Lambert, and Kerner 2016). Interestingly, miR-185 has also been linked to telomere damage and accelerated aging (Li et al. 2020), which is another finding consistently reported for BD (Huang et al. 2018; Fries et al. 2020). All in all, the replication of its upregulation in BD and its link to many BD-related pathways suggest it as a key target in the disorder and warrant further studies focusing on this particular miRNA.

### Limitations

Many limitations of this study need to be acknowledged. The use of EVs enriched for neural origin as a source of biomarkers in clinical studies has been supported by many investigations in the field of neuropsychiatry (Mustapic et al. 2017). Initial studies have proposed that the neural cell adhesion protein L1CAM (CD171) could be a good target for immunoprecipitation of EVs due to its specific expression in neural tissue (Mustapic et al. 2017), which was followed by multiple studies using this marker for the isolation of so-called neural-derived EVs (NDEVs) (Gomes and Witwer 2022). Nonetheless, this approach has been met with controversy over the past few years given evidence that L1CAM is also expressed in other tissues, thus reducing the neural specificity of EVs obtained with this marker (Norman et al. 2021; Hill 2019). Indeed, a recent study found that EV subpopulations can be identified in human biofluids, including of neuronal origin, but further supported that L1CAM may not serve as a reliable marker to isolate NDEVs (Kadam et al. 2024). For this reason, the neuronal specificity of these EVs cannot be guaranteed, although recent work revisiting this controversy provides further support for L1CAM as a reliable biomarker of NDEVs (Nogueras-Ortiz et al, 2024). In addition, the sample sizes for both sub-studies were small, which may have led to underpowered statistical analyses and prevented us from further exploring the results in specific subgroups of individuals. The samples were heterogeneous in terms of medication use, acute mood symptoms, and clinical presentations, all of which may have acted as confounders in our analyses. Many unmeasured external factors could also be influencing the levels of miRNAs measured in our study, such as diet, physical activity, and sleep patterns. Our results should also be acknowledged as preliminary given our lack of validation with alternative, complementary methods (such as quantitative polymerase chain reaction), or with independent, replication samples. Finally, while L1CAM is still a controversial neural-specific marker (as previously discussed), we also lack a more detailed characterization of the EVs isolated with complementary methods, such as Nanoparticle tracking analysis or electron microscopy.

## Conclusion

This is the first study comparing miRNA expression patterns associated with BD in peripheral blood L1CAM and bulk EVs, providing further insight into the possible pathophysiology of BD including a potential window into brain miRNA expression differences *in vivo*. The use of L1CAM EVs as potential liquid biopsies of brain gene expression in BD and other psychiatric disorders is also supported by the established relevance of the identified target genes and pathways known to be relevant to BD’s pathophysiology, such as BDNF and insulin resistance, as well as the association of significant miRNAs with clinical features in the L1CAM EVs analysis only. Further investigations will be required to better understand the functional implications of the observed miRNA alterations, as well as their concordance with studies conducted in post-mortem brain tissues. In particular, given the higher burden for certain diseases (e.g., diabetes) in individuals with BD, miRNA alterations may be one possible pathway to disease risk, although this hypothesis is preliminary. Regardless, findings offer a promising first step to incorporating brain-specific miRNAs into monitoring as well as intervention work.

## Supporting information

Supplementary Results

## Data Availability

Raw RNA-sequencing data and accompanying phenotype information are deposited at the NIMH Data Archive (NDA, collection ID #3300).

## Acknowledgments

We would like to thank the study participants for their willingness to participate in the study.

## Author contributions

Design and conceptualization of the study, funding acquisition: GRF and JQ. Sample collection: JCS, GS, and JQ. Data generation and processing: GRF, SM, CNCL, and WZ. Data analysis: JW, SM, and MCT. Scientific discussion and interpretation of results: GRF, JQ, CNCL, WZ, MCT, GS, JQ. Writing – original draft: GRF, SM, JQ. Writing – review and editing: GRF, SM, GS, JCS, JQ.

