## Supplementary Results for "Differential microRNA expression profiling of peripheral blood L1CAM neural-enriched and bulk extracellular vesicles in individuals with bipolar disorder"

**Supplementary Table 1.** Overlapping microRNAs between the analysis of L1CAM (Study 1) and bulk (Study 2) extracellular vesicles.

| **miRNA** | **log2FC (Study 1)** | **FDR Q (Study 1)** | **log2FC**  **(Study 2)** | **FDR Q**  **(Study 2)** |
| --- | --- | --- | --- | --- |
| hsa-miR-486-5p | -1.4669665 | 9.73E-05 | 1.00710602 | 0.02131364 |
| hsa-miR-140-3p | -2.1562699 | 0.00679948 | 0.74282238 | 0.04412134 |
| hsa-miR-1-3p | 2.32711194 | 0.09897149 | 2.03756803 | 0.06381652 |
| hsa-miR-486-3p | -1.4651403 | 9.73E-05 | 1.75176683 | 0.09881026 |

FC – fold change; FDR – false discovery rate.

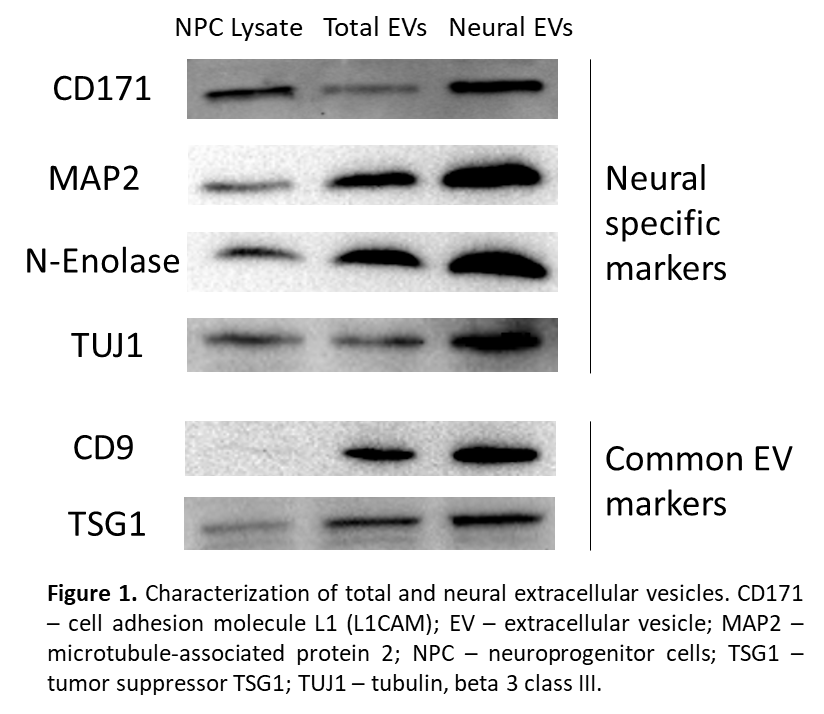
**Supplementary Figure 1.** Characterization of bulk and L1CAM (neural-enriched) extracellular vesicles. CD171 – cell adhesion molecule L1 (L1CAM); EV – extracellular vesicle; MAP2 – microtubule-associated protein 2; NPC – neuroprogenitor cells; TSG1 – tumor suppressor TSG1; TUJ1 – tubulin, beta 2 class III.

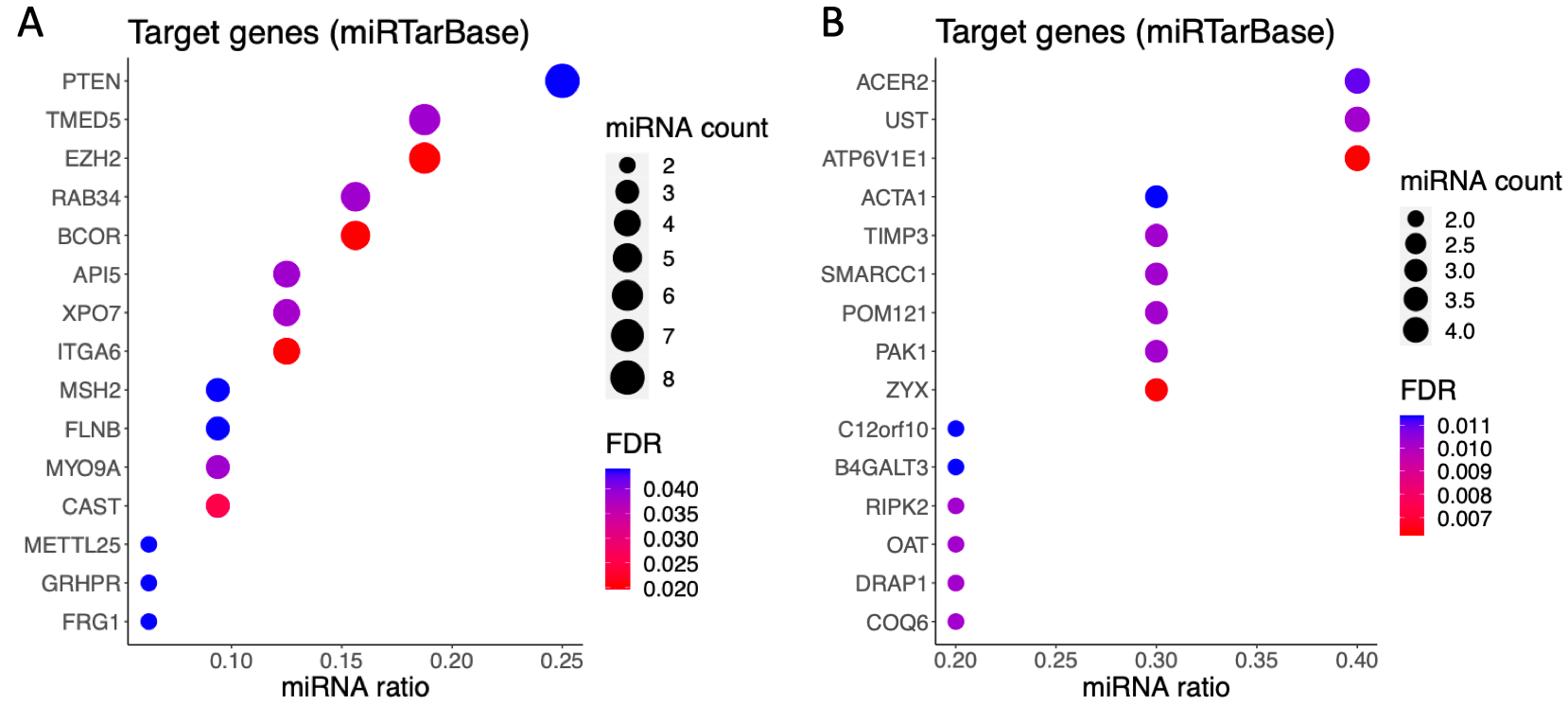

**Supplementary Figure 2.** Target genes of the differentially expressed microRNAs in A) L1CAM (neural-enriched) extracellular vesicles and B) bulk extracellular vesicles (according to miRTarBase).

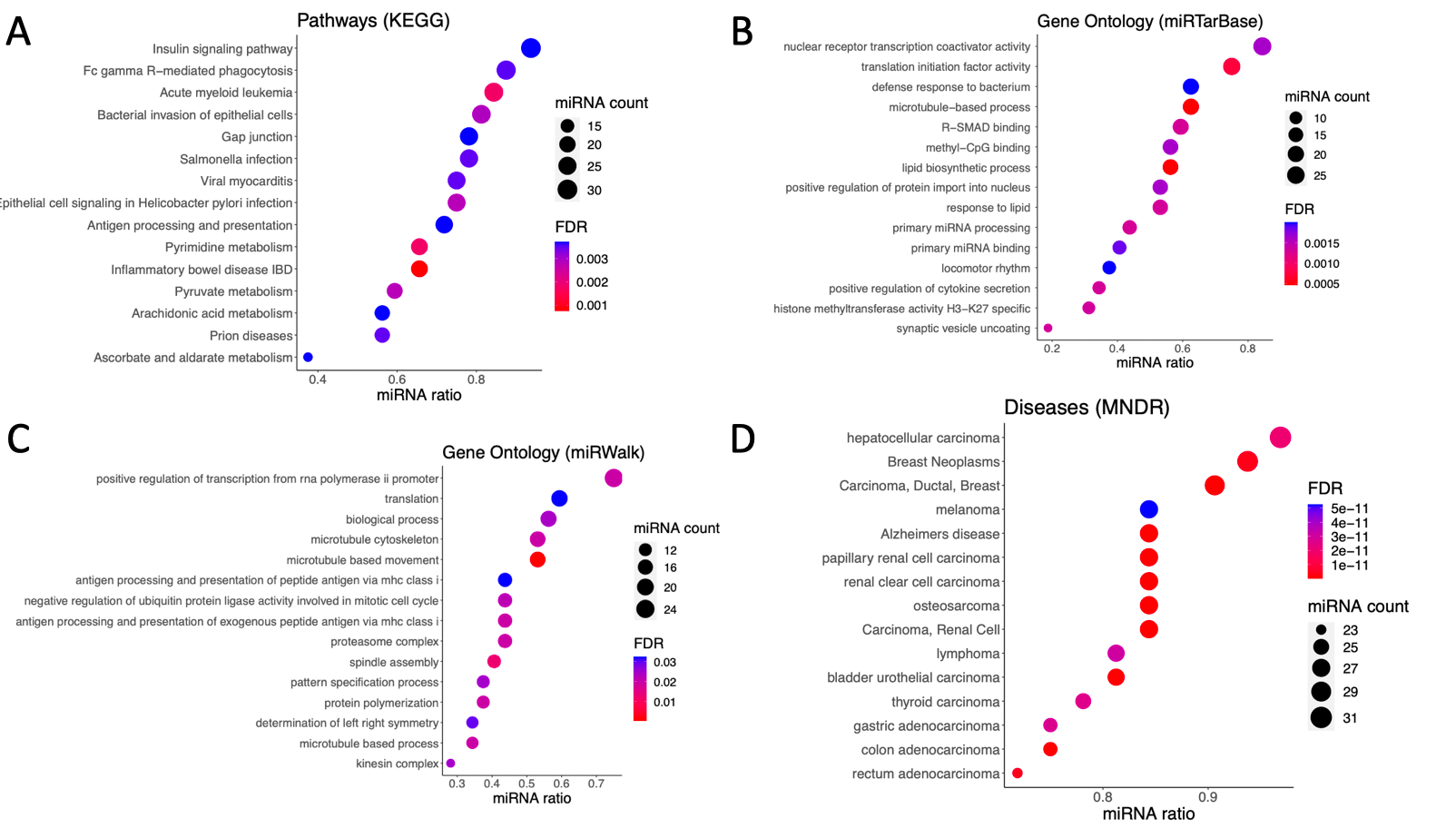

**Supplementary Figure 3.** Pathway analyses with differentially expressed microRNAs in L1CAM (neural-enriched) extracellular vesicles. A) Pathways identified with KEGG database; B) gene ontology terms identified with miRTarBase; C) gene ontology terms identified with miRWalk; d) disease terms identified by the mammal ncRNA–disease repository (MNDR).

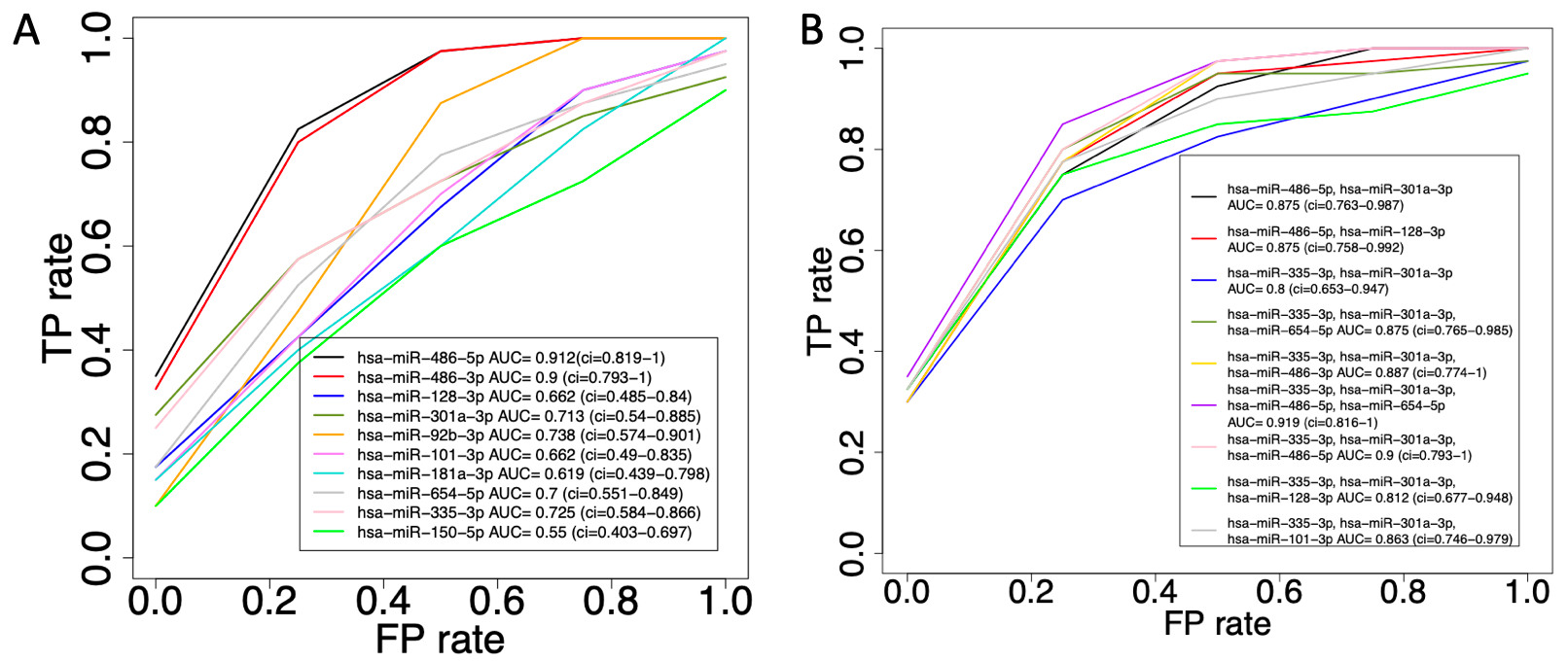

**Supplementary Figure 4.** Receiving operating curves (ROC) for individual (top-ranked) microRNAs identified to be differentially expressed between individuals with bipolar disorder and controls in L1CAM (neural-enriched) extracellular vesicles. AUC - area under the curve; ci - confidence interval, FP - false positive; TP - true positive.

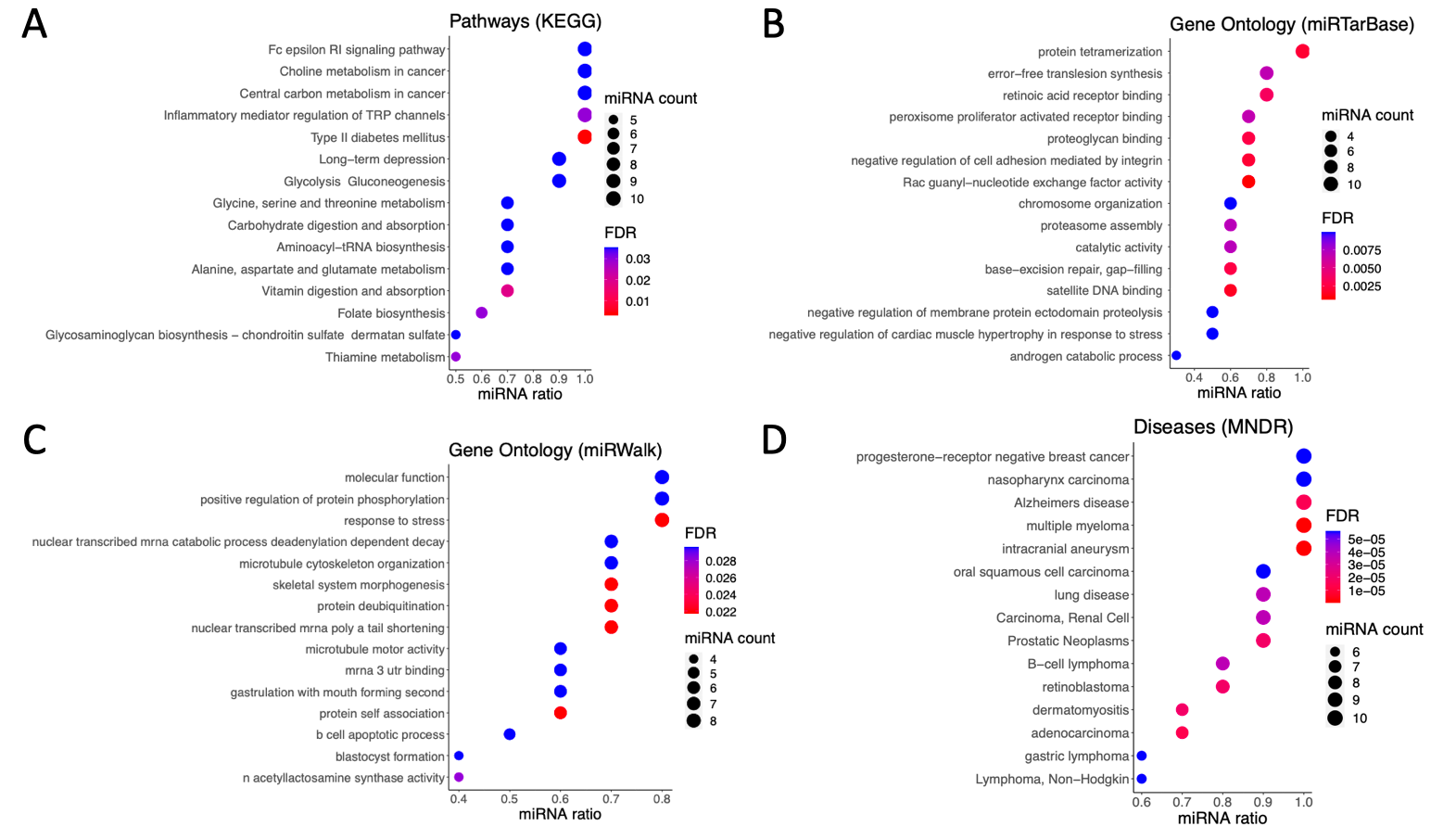

**Supplementary Figure 5.** Pathway analyses with differentially expressed microRNAs in bulk extracellular vesicles. A) Pathways identified with KEGG database; B) gene ontology terms identified with miRTarBase; C) gene ontology terms identified with miRWalk; d) disease terms identified by the mammal ncRNA–disease repository (MNDR).

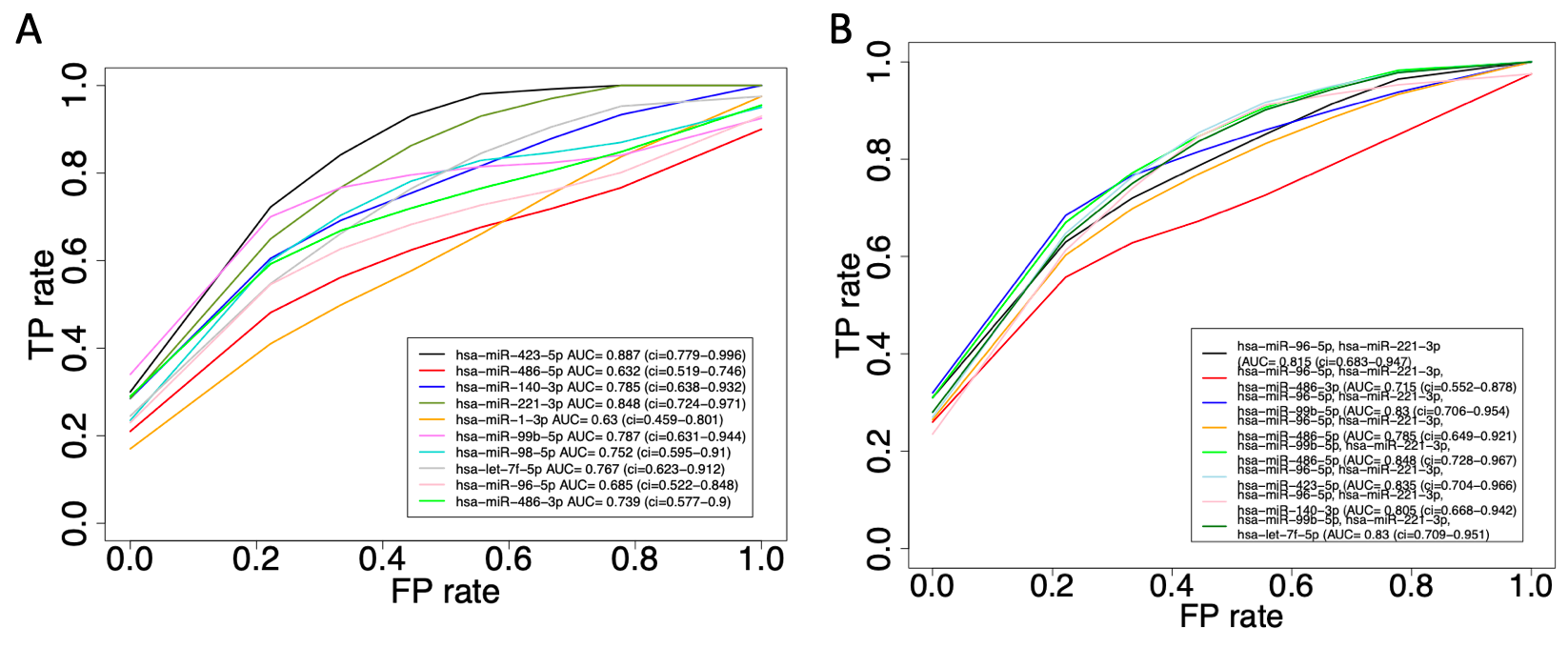

**Supplementary Figure 6.** Receiving operating curves (ROC) for individual (top-ranked) microRNAs identified to be differentially expressed between individuals with bipolar disorder and controls in bulk extracellular vesicles. AUC - area under the curve; ci - confidence interval, FP - false positive; TP - true positive.
